# A Cross Sectional Study to Assess Perception and Behavior towards COVID-19 Vaccine Among Students and Faculties of Nursing Colleges at Anand and Kheda Districts, Gujarat

**DOI:** 10.1101/2021.07.18.21260710

**Authors:** Kailash Nagar, Christian Selina, Parmar Anushka, Patel Devanshi, Bhatt Dhruti, Dalwadi Jinal, Brahmbhatt Manjari, Hiral Janki

## Abstract

**Introduction:** On 11 March 2020, World Health Organization (WHO) announce that COVID-19 as a pandemic health problem [1,2]. According to report on 25 January 2021, worldwide cases reached to 100 million [3]. The first confirmed case of COVID-19 in India was reported on 30, January 2020 in Kerala and affected individual had come from Wuhan, china [4]. Corona virus is spread over 220 countries across the world [2]. In order to prevent corona virus all the countries are taken initiative to prepare effective vaccine against corona virus. There is not any effective treatment is available for corona so far vaccine is the key preventive aspect against corona virus. There are presently above 125 vaccines go through pre-clinical investigation for covid-19[5].

India has develop two types of vaccine (Covaxin and covidshield) in primary phase and from 01 January 2021, Indian government started vaccination namely Covidshield to health workers (front line workers) [6]. So in primary stage target set to cover 30 million health workers followed by policeman and old age peoples to be vaccinated against corona [6]. During primary phase of corona vaccine we don’t have appropriate research and literature, about side effects and how far vaccine is reliable that why due so some miner side effect and negative media publicity peoples are very scared to take vaccine. So few peoples were started denial get vaccinated.

**Aims:** The current study is aimed to assess the perception and behavior towards Covid-19 vaccine among students and faculties of nursing colleges those who have taken vaccine against corona. The researcher wan to explore the positivity through the research result to reduce the negative mindset of the peoples toward corona vaccine, Because in India few peoples has fear to take vaccine against corona due to negative media publicity and scared of side effect.

**Objective:**

1. To assess the existing level of perception toward COVID 19 vaccine among students and faculties of Nursing colleges at Anand and Kheda Districts.
2. To assess the behavior towards COVID 19 vaccine among students and faculties of Nursing colleges at Anand and Kheda Districts.
3. To find out the association between selected socio-demographic variables and perceptions towards COVID 19 vaccine.

**Methodology:** *Design and Setting:* Descriptive cross sectional survey research design was used and non-probability (snowball) sampling method was used to drawn samples through online Google form [7]. Due to Covid-19 pandemic situation research has adopted online snowball sampling method, where after tool validation from various subject experts, all questions were plots on Google form and inform consent form also has been conducted online prior to data collection from the samples[8]. Prior to data collection written setting permission obtain from nursing colleges principals, for the data collection researcher were selected total 03 nursing institutes which were namely Dinsha Patel College of Nursing, Vinayaka College of Nursing, Nadiad, Vinayaka College of Nursing, Anand, and Zydus College of nursing, Anand. The total sample size was 254 nursing college students and faculties. The tool consist of following Section-01 Demographic variables, section-02 Nursing students and faculties information related to covid-19 vaccination during 1^st^ and 2^nd^ dose and Section-3 Questions related to perception and behaviour towards COVID 19 vaccine.

*Statistical Analysis used:* Descriptive statistics applied where, data were analyzed by using SPSS software, and Frequency, percentage, tables etc. were used to represent the statistical data in the tables and graph and figure. Chi-square test was used to assess the significant association between the demographic and level of perception to test the hypothesis.

*Results:* Out of 254, 245(96.45%) were belong age 17-25 years, 219(86.22%) were females, 53(20.87%) were study Diploma course and 178(70.08%) were study degree course, 223(87.79%) belong to Hindu, religion, 227(89.37%) were Unmarried, 134(52.75%) were from urban area, and rest 120 (47.24%) belongs to Rural area. Sources of information about COVID 19 vaccine 109(42.92%) got from online media, 44(17.32%) from television, 243(95.67%) received free of cost corona vaccine, 199(78.35%) mindset was not influenced by negative media publicity about vaccine, 248(97.63%) do not have any co-morbidities. 219(86.22%) taken Covid-19 vaccine empty stomach. 221(87%) of samples were taken willingly vaccine, 205(80.71%) of samples were received covidshield vaccine and others 49(19.29%) were taken Covaxin, 109(42.91%) samples noticed mild fever, 53(20.87%) samples noticed moderate fever, 18(7.08%) noticed severe fever and rest 74(29.13%) didn’t noticed fever.

*Conclusions:* Regarding perception and behaviour towards COVID 19 vaccine, the majority of samples has good perception and behaviour, 74.00% has good perception and only 23.00 had moderate to poor perception, majority of participant were willingly taken vaccine and agree to recommend to others, not evidence any serious side effect due to vaccination.

## INTRODUCTION

On 30 January, World Health Organization (WHO) 2020, announce COVID-19 as a public health crisis and afterwards On 11 March 2020, World Health Organization (WHO) announce the corona virus disease 2019 a pandemic (COVID-19) [1].

Vaccination was one of the greatest cost-efficient, inhibitory actions [9]. Vaccines were the upmost essential public wellness actions and highly successful method to save public from covid-19 [10]. The world is presently working for the quick evolution of the COVID 19 vaccine. A successful COVID 19 vaccine should be useful, effective, set free from any side effect and affordable for local people in the world [11,14].

There are presently above 125 vaccines go through pre-clinical investigation for covid-19. The vaccines are than go into three phases of clinical tests, India has already rolled out a huge coronavirus effort to utilize two vaccines, Covishield and Covaxin [5,16].

The covid-19 vaccine was introduced on 16^th^ January, 2021. Health personnel and frontline workers were the first group who get the opportunity to get COVID-19 vaccine and after them individuals who are above 50 years of age and individuals who are under 50 years and suffering from co-morbidity conditions were the second group for COVID-19 vaccination [6].

There were two doses of covid-19 vaccine which would be offered in 28 days’ gap. The efficiency of vaccine starts later on 14 days of taking the second dose. The covid-19 vaccine was extremely fruitful against covid-19[11]. some experts declare that the vaccine protected against covid-19 in 62% of those who received two full doses and 90% of those who initially received half dose [12,13,15].

## MATERIALS AND METHODS

### Research Approach

Non Experimental, Descriptive survey approach

### Research Design

Cross sectional survey.

### Research Variables

1. **Dependant variables:** Perception and behavior toward covid-19 vaccine
2. **Demographic variables:** demographic variables of Nursing Student’s such as Age, Gender, Course, Year, Marital status, vaccination history, side effects.

### Sampling method

The E-survey was prepared online and hyperlink of the survey was distributed to students using mobile group messaging application. It was made sure in a class that most of the students are having smart mobile devices and sufficient Internet connectivity to fill up the form online. Students who were not using Internet were encouraged to take help from their friends having Internet enabled device. Prior to the distribution, students were made clear about the objectives of this study and inform consent form. It is to be noted that student participation was voluntarily and they could opted not to fill up the E-survey.

### Instrument for Data Collection

For the data collection toll has been prepared in three categories. 1. Questionnaire related to Covid-19 vaccine 1^st^ dose, 2. Questionnaire related to Covid-19 vaccine 2^nd^ dose, and 3 point likert scale to assess the perception and behavior.

#### Study population

Nursing College Students. And Faculties.

#### Study Sample

Nursing students and faculties who received covid-19 vaccine

#### Study Setting

04 nursing institutes of the kheda and Anand District Gujarat.

#### Sample Size

254 Nursing College Student and Faculties.

## SAMPLE CRITERIA

### Inclusion criteria

1. Students and faculties of nursing colleges of both gender of age between 17-60 years.
2. Those who have taken COVID – 19 vaccine.

### Exclusive criteria

1. Those who are not willing to participate in study.
2. Those who have not taken vaccine.

### Tool for Data Collection

Section-I: Consist of Demographic variables.

Section-II: Consist of Questionnaire related to Covid-19 vaccine 1^st^ dose.

Section-III: Consist of Questionnaire related to Covid-19 vaccine 2^nd^ dose.

Section-IV: Consist of 3 point likert scale to assess the perception and behavior.

## RESULTS

### Section I: Demographic variables of nursing students and faculties

The [Table/Fig-1] portrays that majority participants (96%) age below 25 years, majority (86%) were female, (70%) were undergraduate students.

**Table/Fig-1]:**
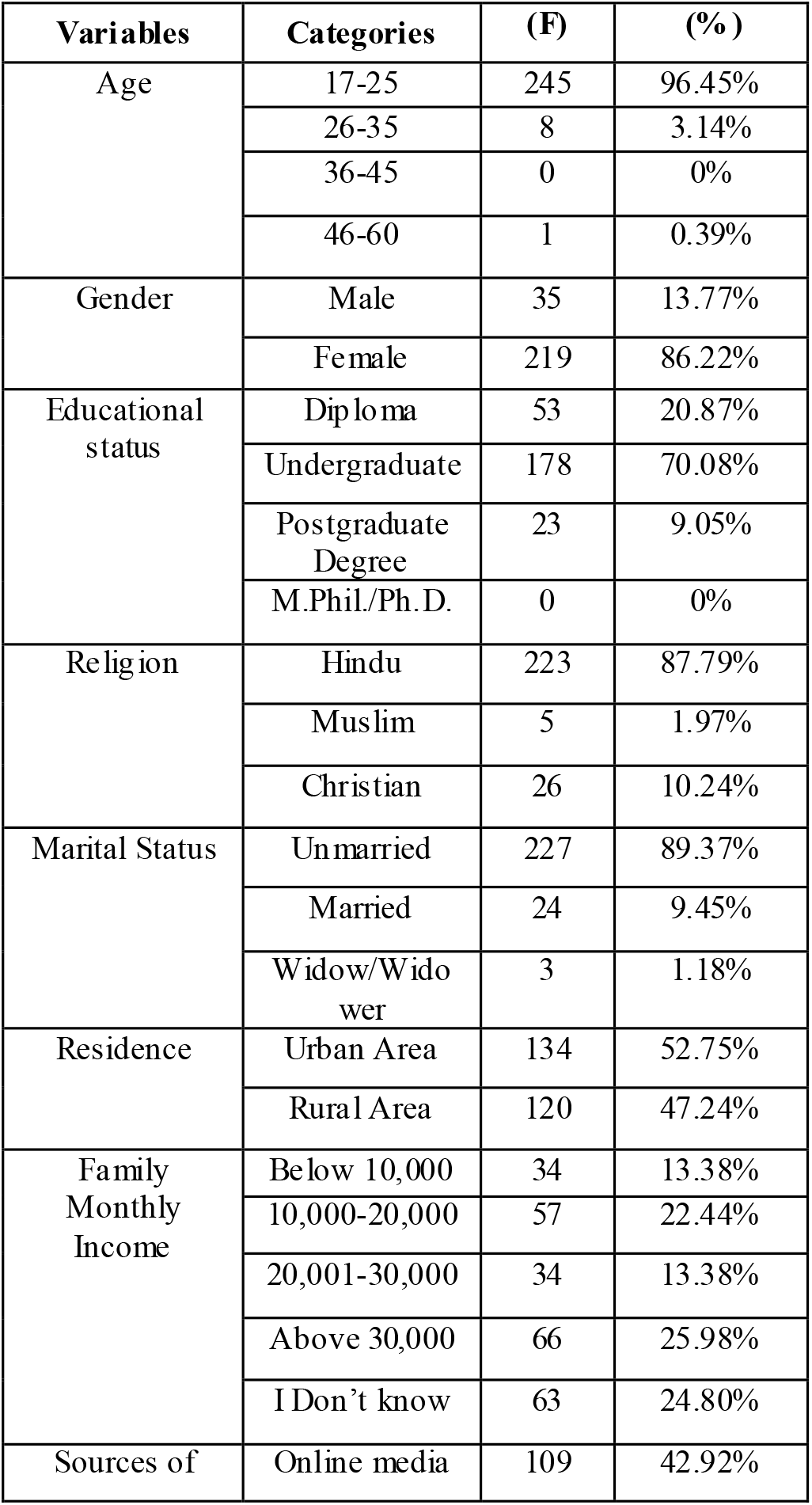
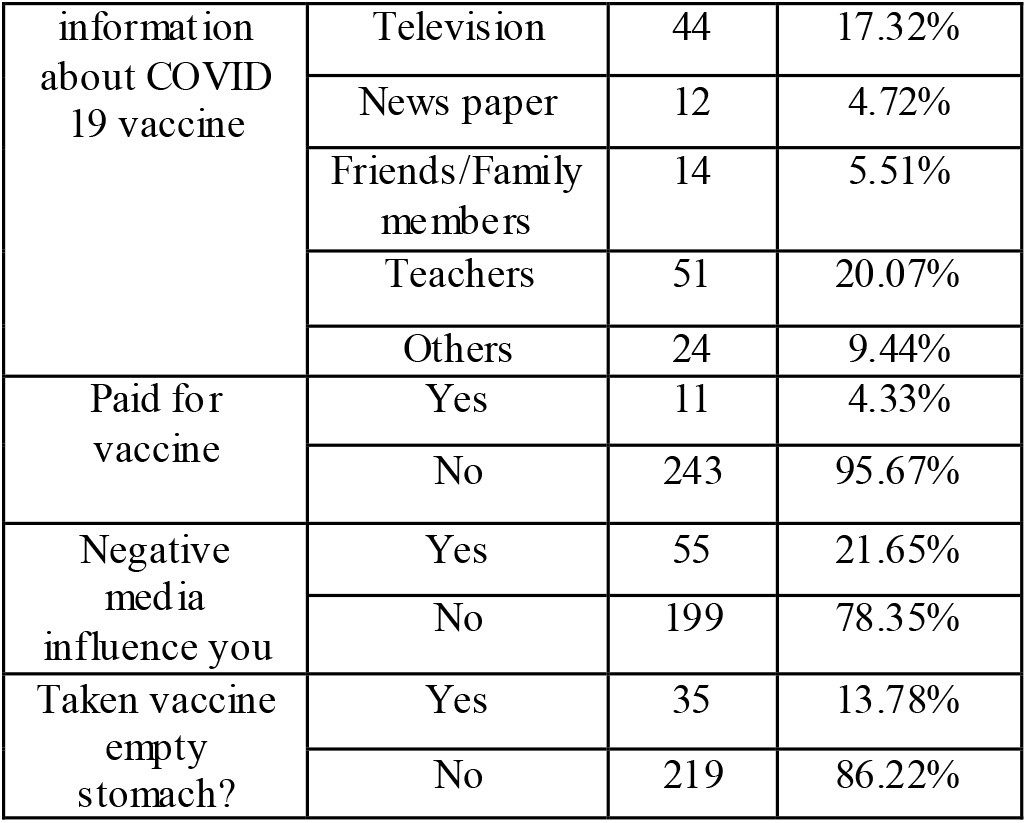
Frequency and percentage distribution according to demographic variables. (N=254) Key:- (F)= Frequency, (%)= Percentage

[Table/Fig-1] depicts majority (87%) were belong to Hindu religion, (89%) were unmarried, (95%) received vaccine free of cost, majority (86%) taken vaccine empty stomach.

### Section II: Distribution according to information during 1^st^ dose of covid-19 vaccine

The [Table/Fig-2] depicts majority (87%) willingly taken vaccine, (80%) were taken Covishield vaccine, (80%) does not have any serious side effects after taken vaccine, (42%) mild fever and last longer for 1-2 days. (43.70%) had mild pain on the vaccine site, 203(79.92%) participant do not have fear prior to take corona vaccine.

**[Table/Fig-2].** Frequency and percentage distribution according to information during 1^st^ dose of covid-19 vaccine (N=254).

| Statement | Total | % |
| --- | --- | --- |
| **Status of COVID-19 vaccination?** |  |  |
| Willingly taken | 221 | 87.00 |
| Not Willingly taken | 33 | 13.00 |
| **Which COVID-19 Vaccine injected in your body?** |  |  |
| Covishield | 205 | 80.7 |
| Covaxin | 49 | 19.3 |
| **Do you have any severe side effects after taking 1<sup>st</sup> dose of COVID-19 vaccine?** |  |  |
| Yes | 51 | 20.0 |
| No | 203 | 80.0 |
| **After taking 1<sup>st</sup> dose of vaccine status of fever?** |  |  |
| Mild Fever | 109 | 42.9 |
| Moderate Fever | 53 | 29.8 |
| Severe Fever | 18 | 7.0 |
| None of above | 74 | 29.1 |
| **Duration of Fever after taking 1<sup>st</sup> dose of COVID-19 vaccine?** |  |  |
| 1-day | 97 | 38.1 |
| 2-days | 79 | 31.1 |
| 3-days or more | 08 | 3.1 |
| None of above | 70 | 27.7 |
| **Did you required hospitalization after taking 1<sup>st</sup> dose of COVID-19 vaccine?** |  |  |
| Yes | 10 | 4.0 |
| No | 244 | 96.0 |
| **Status of injection site pain after taking 1<sup>st</sup> dose of COVID-19 vaccine?** |  |  |
| Mild Pain | 111 | 43.7 |
| Moderate Pain | 65 | 25.5 |
| Severe pain | 19 | 7.5 |
| None of above | 59 | 23.2 |
| **Duration of injection site pain after taking 1<sup>st</sup> dose of COVID-19 vaccine?** |  |  |
| 1-day | 67 | 26.4 |
| 2-days | 68 | 26.8 |
| 3-days or more | 60 | 23.6 |
| None of above | 59 | 23.2 |
| **After taking 1<sup>st</sup> dose of COVID-19 vaccine which other side effects do you have?** |  |  |
| Nausea and vomiting | 05 | 2.0 |
| Lethargy | 18 | 7.09 |
| Headache and Body ache | 131 | 51.1 |
| None of above | 100 | 39.0 |
| **Do you have fear prior to take COVID-19 vaccine?** |  |  |
| Yes | 51 | 20.0 |
| No | 203 | 80.0 |

### Section III: Distribution according to information during 2^nd^ dose of covid-19 vaccine

[Table/Fig-3] depicts majority (89%) willingly taken vaccine, (90%) does not have fear (97%) does not have any serious side effects after taken vaccine, (37%) mild fever and last longer for 1-2 days. (44.70%) had mild pain on the vaccine site.

**[Table/Fig-3].**
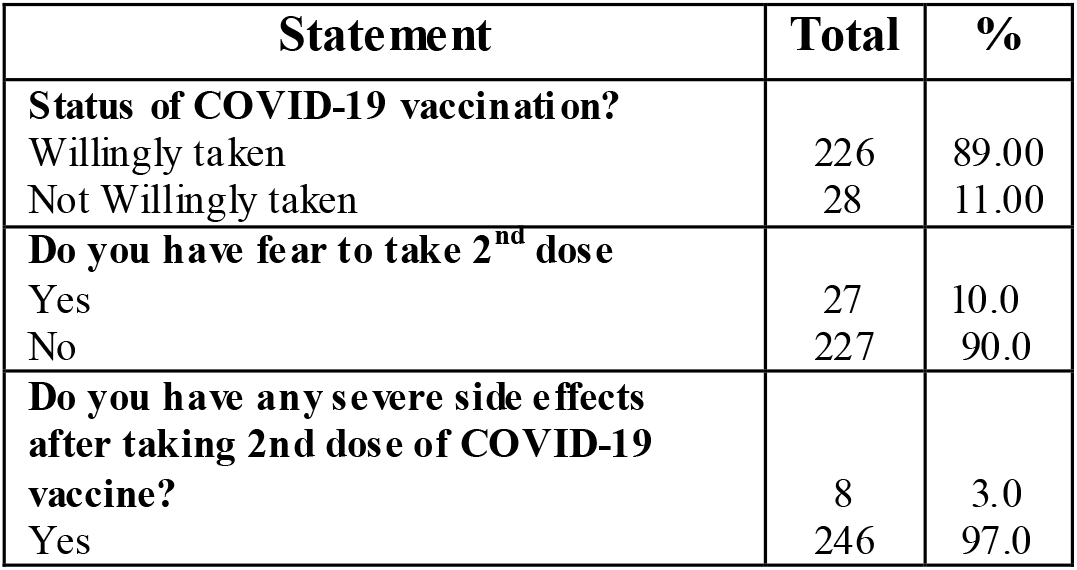
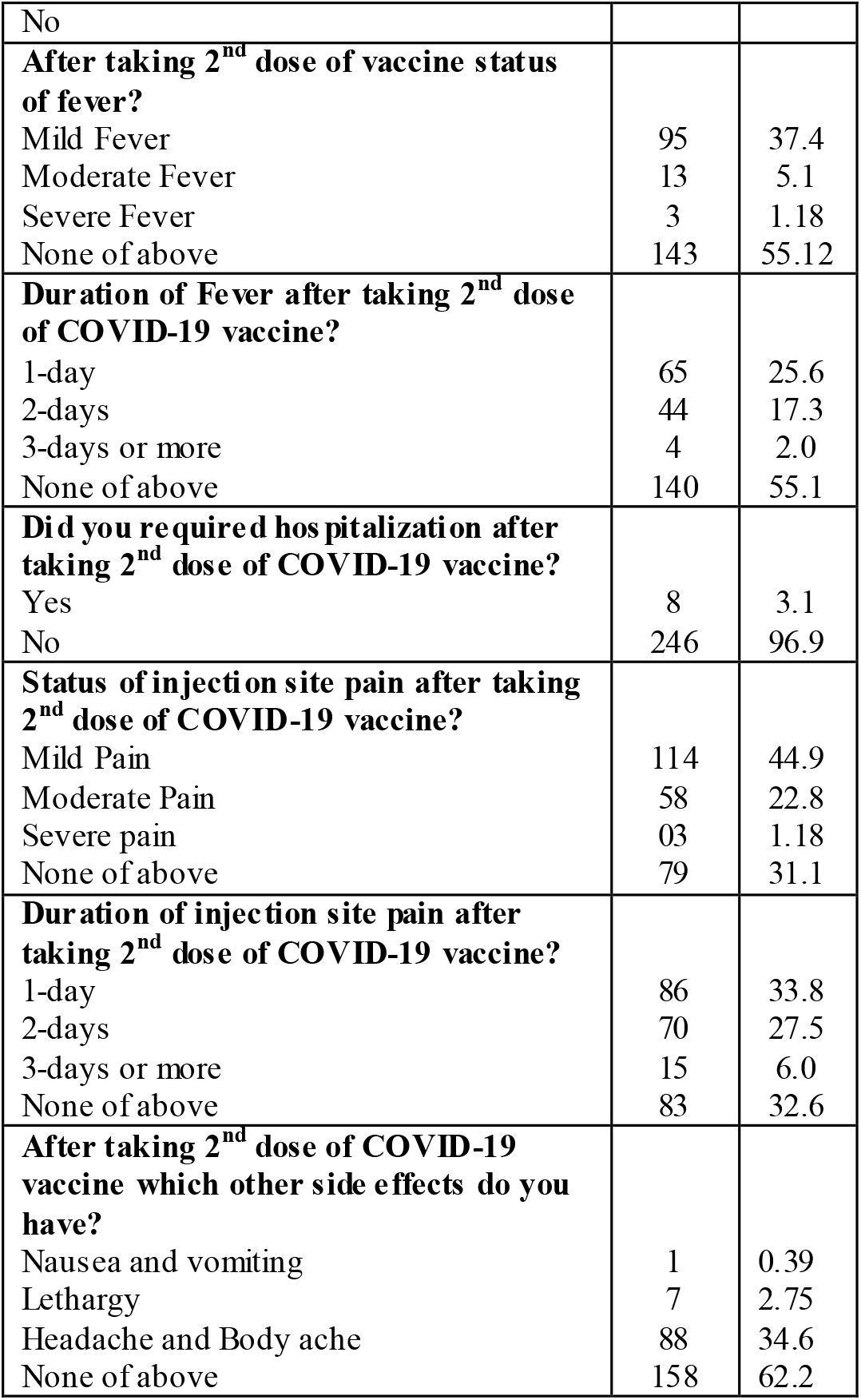
Frequency and percentage distribution according to information during 2^nd^ dose of covid-19 vaccine (N=254).

### Section IV: Distribution according to Perception and behavior toward covid-19 vaccine

[Table/Fig-4] depicts majority only 7(2.7%) had poor perception, 60(23.7%) had moderate perception, 187(73.6) majority of participant had good perception.

**[Table/Fig-4].** Frequency and percentage distribution according Perception and behavior toward covid-19 vaccine.

| Perception and behavior level | Frequency | Percentage |
| --- | --- | --- |
| Poor Perception | 7 | 2.7 |
| Moderate Perception | 60 | 23.7 |
| Good Perception | 187 | 73.6 |
| **Total** | **254** | **100%** |

**[Table/Fig-5].** Range, Mean, Standard deviation, Median score of perception and behavior among nursing students & Faculties. (n=254)

| Max. Score | Perception and behavior Score |  |  |  |  |
| --- | --- | --- | --- | --- | --- |
|  | Min-Mx | Range | Mean | SD | Median |
| 60 | 23-60 | 37 | 48.68 | 7.32 | 50.50 |

### Section V: Distribution according to Association between perception and selected demographic variables

The [Table/Fig-7] depicts outcome of chi-square test results, In reference to the association of perception and behaviour with selected demographic variables, there was significant association of perception with sources of corona vaccine information and rest of variable found not significant, at 0.05 level of significant.

**[Table/Fig-6].** Level of Perception and behavior toward covid-19 vaccine likert scale (n=254)

| Statement | A | U | DA |
| --- | --- | --- | --- |
|  | Percentage |  |  |
| Will you recommend COVID 19 vaccine to others? | 74.8 | 10.2 | 19.9 |
| Getting myself vaccinated for COVID 19, would be good way to protect myself against infection? | 81.4 | 10.2 | 8.2 |
| Does COVID 19 symptoms after taken vaccine may differ from one person to another? | 41.7 | 26.3 | 31.8 |
| Do you think COVID 19 vaccination is an effective way to prevent and control COVID 19? | 74.8 | 12.5 | 12.5 |
| Is Corona infection providing better immunity than COVID 19 vaccine? | 44.8 | 24.0 | 31.1 |
| Is COVID 19 vaccine safe for all? | 56.2 | 19.2 | 24.4 |
| Do you think all has to take COVID 19 vaccine? | 68.1 | 16.9 | 14.9 |
| COVID 19 vaccine may be fatal? | 9.05 | 44.8 | 46.0 |
| COVID 19 vaccine can lead to serious health issues? | 7.8 | 24.8 | 67.3 |
| COVID 19 vaccine is sufficient for preventing Corona virus to all? | 45.2 | 22.8 | 31.8 |
| I believe a vaccine can help control the spread of COVID 19? | 23.6 | 60.6 | 15.7 |
| Is COVID 19 vaccine become available, it should be mandatory for all? | 55.9 | 23.6 | 20.4 |
| Is COVID 19 vaccine will be given to everyone simultaneously? | 56.2 | 28.3 | 15.3 |
| Is it mandatory to take the vaccine? | 50 | 27.9 | 22.0 |
| Is it necessary for a COVID recovered person to take the vaccine? | 57.0 | 27.5 | 15.3 |
| The vaccine introduced in India be as effective as the ones introduced in other countries? | 53.9 | 35.8 | 10.2 |
| Does one need to follow preventive measures such as wearing mask, hand sanitization, social distancing after receiving the COVID 19 vaccine? | 72.4 | 15.7 | 11.8 |
| Are there any common side effects of this vaccine? | 53.1 | 14.5 | 32.2 |
| Does negative media information's work as a barrier in COVID 19 vaccination? | 47.6 | 25.1 | 27.1 |
| Will you follow all the COVID 19 protocols once you get vaccinated? | 73.6 | 17.3 | 9.0 |

**[Table/Fig-7].** outcome of chi-square test results to find significant association between selected demographic variables of nursing students and faculties (n=254)

| Variables | Categories | (f) | Chi-Square | Sig. P-value |
| --- | --- | --- | --- | --- |
| Age | 17-25 | 245 | 4.063<br>df=9 | 0.907<br>NS |
|  | 26-35 | 8 |  |  |
|  | 36-45 | 0 |  |  |
|  | 46-60 | 1 |  |  |
| Gender | Male | 35 | 0.916<br>Df=9 | 1.00<br>NS |
|  | Female | 219 |  |  |
| Educational status | Diploma | 53 | 5.89<br>df=9 | 0.750<br>NS |
|  | Undergraduate | 178 |  |  |
|  | Postgraduate Degree | 23 |  |  |
|  | M.Phil./Ph.D. | 0 |  |  |
| Religion | Hindu | 223 | 5.278<br>Df=9 | 0.908<br>NS |
|  | Muslim | 5 |  |  |
|  | Christian | 26 |  |  |
| Marital Status | Unmarried | 227 | 1.891<br>df=9 | 0.993<br>NS |
|  | Married | 24 |  |  |
|  | Widow | 3 |  |  |
| Residence | Urban Area | 134 | 4.046<br>df=6 | 0.67<br>NS |
|  | Rural Area | 120 |  |  |
| Family Monthly Income | Below 10,000 | 34 | 19.86<br>df=15 | 0.177 |
|  | 10,000-20,000 | 57 |  |  |
|  | 20,001-30,000 | 34 |  |  |
|  | Above 30,000 | 66 |  |  |
|  | I Don't know | 63 |  |  |
| Sources of information about COVID 19 vaccine | Online media | 109 | 32.87<br>df=18 | 0.18<br>Significant |
|  | Television | 44 |  |  |
|  | News paper | 12 |  |  |
|  | Friends/Family members | 14 |  |  |
|  | Teachers | 51 |  |  |
|  | Others | 24 |  |  |

## Conclusion

On the basis of findings of this study the following conclusion were drawn:

The purpose of the present study is to assess the attitude regarding online lecture after the impact of COVID-19 at selected nursing college Nadiad. The study consisted of 136 samples that were selected on the basis of simple randomization techniques. Based on the objective, the data analysis was done by calculating the mean, percentage, standard deviation. Result revealed that an attitude of nursing students towards Regarding the choice of gadgets to attends online class of students out of 136 samples, 132(97.05%) was mobile, 2(1.47%) was laptop2(1.47%). Regarding the network quality of students out of 136 samples, 33(24.26%) was poor, 79(58.08%) was average, 24 (17.64%) good. Revealed that the distribution of sample according to Attitude regarding Online Classes were 7 (5.14%) had Inadequate Attitude 86(63.25%) had Moderate Attitude, 43(31.61%) Had adequate attitude.

## Data Availability

all important data are included in the manuscript

## Conflict of Interest

Nil (there is not any conflict of interest between the all authors)

## Source of Funding

Self (Contributed by all authors)

## Ethical Clearance

The study was approved by the institutional ethical committee of Dinsha Patel college of nursing, research committee, There are total 15 members in the committee from various field. **The ethical approval reference number is DPCN/2**^**nd**^**IEC/2020-21/13** and a formal written permission was gathered from the authority of or Principal of Institute prior to data collection

## Statement of Informed consent

Yes informed consent form was acquired from the participants prior to data collection.

## Acknowledgement

Special thanks to all the participants of the study and principals of the selected colleges for provide us permission for data collection.

## Notes

### Competing Interest Statement

The authors have declared no competing interest.

### Clinical Trial

Clinical trial was not obtain and its not required because its an descriptive study and not any harm to the participants of the study, the survey data has been collected online from E-survey tool

### Funding Statement

self funded by all the authors

### Author Declarations

Ethical Clearance: The study was approved by the institutional ethical committee of Dinsha Patel college of nursing, research committee, There are total 15 members in the committee from various field. The ethical approval reference number is DPCN/2ndIEC/2020-21/13 and a formal written permission was gathered from the authority of or Principal of Institute prior to data collection. participant consent form was obtain before data collection and prior to become pat of the study, the objective and aims of the study were informed to all the participants.

